# Serum fibrinogen level and fibrinogen administration in patients with traumatic brain injury: a systematic review and meta-analysis protocol

**DOI:** 10.1101/2024.08.26.24312610

**Authors:** Joanne Igoli, Jeremiah Oluwatomi Itodo Daniel, Halleluyah Oludele, Adedoyin Esther Alao, Idemudia Stephen Ogedegbe, Adewale Olaniyan, Michael Adeshola Adebayo, Damilola Matthew, Temidayo Elizabeth Oyepitan, Daniel Brabi, Olatomiwa Olukoya, Temidayo Osunronbi

**Author notes:** **Please address all correspondence to:** Dr Joanne Igoli, Deanery of Clinical Sciences, The University of Edinburgh, Edinburgh, United Kingdom., EH16 4SB. Joint-first authors: contributed equally (Joanne Igoli, Jeremiah Oluwatomi Itodo Daniel).

## Abstract

**Introduction:** Traumatic Brain Injury (TBI) is a leading cause of disability and death globally. It has a significant economic burden. Coagulopathy has been identified as one of the key factors contributing to the poor outcomes observed in TBI patients, and it has been theorised that the management of coagulopathy will improve patient outcomes. Low serum fibrinogen levels denote a coagulopathic state, and the therapeutic administration of fibrinogen has been proposed to correct this state. However, there is no consensus on its efficacy in patients with TBI.

This systematic review and meta-analysis seeks to ascertain the prognostic value of serum fibrinogen levels in patients with TBI and assess the effect of fibrinogen administration on these patients.

**Methods:** Using the Preferred Reporting Items for Systematic Review and Meta-Analysis Protocols (PRISMA-P) guidelines, we will perform a comprehensive search of Scopus, Medline, Embase and Cochrane Library to retrieve all original articles that investigate the prognostic value of fibrinogen levels and/or the effect of fibrinogen administration in TBI patients. Primary outcomes include functional outcome and mortality assessments such as the Glasgow Outcome Score and modified Rankin Score. Secondary outcomes include progressive intracranial haemorrhage/contusion and need for surgical intervention. Data collected will encompass participant demographics, measured fibrinogen levels, dose of fibrinogen administered and specified outcome measures.

**Conclusion:** Findings from this study on if fibrinogen level has prognostic value and if fibrinogen administration improves patient outcomes, will help inform future TBI management and shared decision-making. Thus, TBI patient outcomes can be optimised accordingly.

**PROSPERO Registration Number:** CRD42024556497. Available from: https://www.crd.york.ac.uk/prospero/display_record.php?ID=CRD42024556497

## Introduction

Traumatic Brain Injury (TBI) is a leading cause of disability and death, with an incidence of approximately 69 million people per year globally[1] and a prevalence of approximately 75 million [1,2]. The estimated global economic cost of TBI is $400 billion [3]. Approximately half of TBI patients seen in hospitals fail to regain pre-TBI baseline health six months after the injury [2]. Sequelae of TBI range from transient loss of consciousness and confusion to seizures, persistent cognitive decline, coma, and death [4–6].

Coagulopathy in TBI influences the development of intracranial haemorrhage (ICH), leading to poor patient outcomes, including a higher incidence of organ failure, poor neurological function recovery, significant disability, and death [7–9]. Fibrinogen plays a vital role in the clotting cascade of stabilising blood clots. A low fibrinogen level is a marker of coagulopathy. Serum fibrinogen level has been shown to decrease significantly hours after trauma and reaches severely low levels in uncontrolled post-traumatic bleed[10].

Furthermore, adverse associations of low fibrinogen levels in patients with TBI have been shown, such as loss of independence due to severe disability, vegetative state, and death [9,11,12]. It has been postulated that low fibrinogen levels can be used to prognosticate patients with TBI, and therapeutic administration of fibrinogen could improve coagulopathy in these patients, improving their outcomes.

This systematic review aims first to assess the evidence on the prognostic value of fibrinogen in TBI, which would benefit risk-benefit shared decision-making involving healthcare providers, patients, and their families. Secondly, the authors aim to evaluate the evidence on the benefits, if any, of fibrinogen administration in patients with TBI, which, if demonstrated, would transform and broaden treatment options in this group of patients.

## Materials and Methods

### Research Questions

Question 1: Is there a prognostic value of fibrinogen level in patients with traumatic brain injury?

*Population*: Patients with traumatic brain injury (TBI)

*Intervention*: Serum fibrinogen level

*Control*: N/A

*Outcomes*: Glasgow outcome score; need for surgical intervention; mortality; functional outcomes; progressive intracranial haemorrhage/contusion; other.

Question 2: What is the effect of fibrinogen administration on outcomes in patients with traumatic brain injury?

*Population*: Patients with traumatic brain injury (TBI)

*Intervention*: Fibrinogen administered

*Control*: Fibrinogen not administered

*Outcomes*: Glasgow outcome score; need for surgical intervention; mortality; functional outcomes; progressive intracranial haemorrhage/contusion; other.

## Data Collection

### Literature Search Strategy

We searched the abstracts and titles of articles in Medline, SCOPUS, Cochrane, and EMBASE databases from inception to June 01, 2024, using the following search strategy:

#1 head OR craniu* OR crania* OR cerebra* OR cerebru* OR brain* OR forebrain* OR skull* OR hemispher* OR intracran* OR intracerebral

#2 injur* OR trauma* OR damag* OR wound* OR fracture* OR contusion* OR haematoma OR hematoma OR haemorrhage OR hemorrhage

#3 Factor I OR Fibrinogen OR Fibrin OR Fibrinogen concentrate OR Hypofibrinogenemia OR Hypofibrinogenaemia OR Fibrin degradation product OR Coagulopathy OR Clot* OR Haematolog* OR Hematolog* OR Cryoprecipitate OR Riastap OR Fresh Frozen Plasma

#4 #1 AND #2 AND #3

The initial results of the search strategy are shown in Table 1.

**Table 1:**
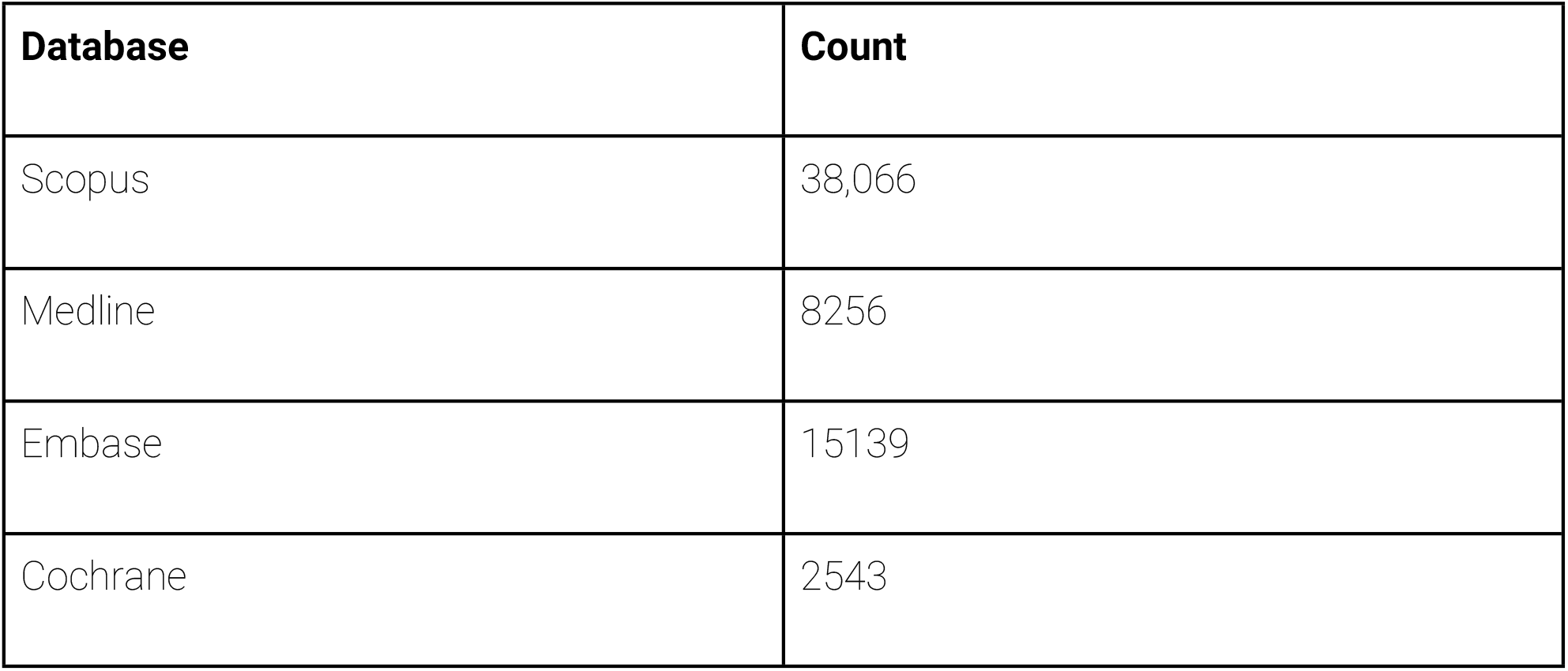
Search Strategy Table.

| Database | Count |
| --- | --- |
| Scopus | 38,066 |
| Medline | 8256 |
| Embase | 15139 |
| Cochrane | 2543 |

## Eligibility Criteria

### Inclusion Criteria

We will include full-text original articles that investigate the prognostic value of serum fibrinogen level and/or investigate the effects of fibrinogen administration on outcomes in TBI patients.

### Exclusion Criteria

Review articles, expert opinions, editorials, letters, conference abstracts, case reports, case series, non-English language papers, paediatric-focussed studies and non-human studies will be excluded. Studies in which fibrinogen was used solely as a marker of disease severity and/or solely for diagnostic reasons will also be excluded.

## Data Extraction

Zotero (Corporation for Digital Scholarship, Virginia, USA) will be used to exclude literature search duplicates. Using Rayyan (Rayyan, Cambridge, Massachusetts, USA), titles/abstracts will be independently screened by two reviewers for inclusion. Eligible full-text articles will be screened further, and subsequent data extracted using the same data extraction proforma on Google Sheets (Google LLC, Mountain View, California, USA). A third reviewer will adjudicate any conflicts in title/abstract screening, full-text screening, and data extraction. The process will be illustrated using a Preferred Reporting Items for Systematic Reviews and Meta-Analyses (PRISMA) [13] flow diagram.

### Data to be extracted

- Title
- Author
- Publication year
- Study design
- Country of study: Based on where the study took place. The first author’s country will be used when the study location is not specified.
- Cause of TBI, if included, such as blunt trauma, penetrating trauma, other
- Neurosurgical procedures carried out such as decompressive craniectomy or craniotomy for extradural clot evacuation.
- Exclusion criteria
- Number of patients
- Age (mean ± SD/median (IQR))
- Sex (male/female proportion)
- Day serum fibrinogen blood sample was obtained
- Average fibrinogen level
- Level of serum fibrinogen that triggered fibrinogen administration (if serum fibrinogen was checked pre-fibrinogen administration)
- Day fibrinogen was administered: Preoperative or postoperative. If postoperative, the day(s) fibrinogen was administered will be extracted
- Dose(s) of fibrinogen administered
- Number of times fibrinogen was administered and the triggers for subsequent administration
- Outcome measure: The nature of the outcome measure to be identified is uncertain. Therefore, there will be no prior criteria. It is anticipated that outcomes such as the following will be identified and used as inclusion criteria:
  ∘ Glasgow Outcome Score (GOS)/Glasgow Outcome Score Extended (GOSE)
  ∘ Modified Rankin Score
  ∘ Mortality
  ∘ Quality of life
  ∘ Progressive intracranial haemorrhage/contusion
  ∘ Other For each outcome measure, the following will be extracted:
  ∘ Length of stay (LOS)
  ∘ Number of patients with event/number without event
  ∘ Length of follow-up
  ∘ Number of patients lost to follow-up
  ∘ Low serum fibrinogen threshold
  ∘ The method used for threshold selection
  ∘ Area under the curve (AUC)
  ∘ Sensitivity
  ∘ Specificity
  ∘ Event ratio with 95% confidence interval (CI)
  ∘ Conclusion (on the association between fibrinogen level and outcome measure; on association between fibrinogen administration and outcome measure).

Multivariate data will be preferred if both multivariate and univariate data are provided. However, univariate data will be used instead in the absence of multivariate data. For AUC, solely univariate data will be used.

## Data Analysis

The direction of effect for the association between serum fibrinogen level or fibrinogen administration and patient outcomes will be categorised as significantly positive, significantly negative, or not statistically significant from articles where the event ratio is reported. Where the area under the curve (AUC) is reported, the effect direction will be grouped as failed (AUC = 0.5 to 0.6), poor (AUC = 0.6 to 0.7), acceptable (AUC = 0.7 to 0.8), good (AUC = 0.8 to 0.9) and excellent (AUC > 0.9) [14]. Dose and response analyses will be done [15], where possible, to evaluate the most effective dose of fibrinogen administration. Side effect frequency and severity will be reported.

Where possible, if ≥5 appropriate articles, meta-analysis will be performed using Review Manager (RevMan) (Copenhagen: The Nordic Cochrane Centre, The Cochrane Collaboration) as follows: each outcome measure will have a pooled extracted odds ratio (OR)/hazard ratio (HR) with the corresponding 95% confidence interval (CI). The generic inverse-variance method will be used to assess the association between fibrinogen level/fibrinogen administration and the dichotomous outcome measure. The Cochrane Q-statistic and I^2^ statistic tests will be used to assess heterogeneity among included studies. Study heterogeneity will be addressed using a random-effects model. Subgroup analyses will be carried out for studies that explicitly state that their study participants had isolated head injuries. Subgroup analyses will be conducted through sensitivity analysis, where the primary analysis is repeated after excluding studies with a high risk of bias. The risk of bias (RoB) in the eligible full-text articles will be assessed by two independent reviewers, with a third reviewer adjudicating any discrepancies.

The RoB in studies investigating the prognostic utility of serum fibrinogen level will be assessed using the Quality in prognosis studies (QUIPs) tool [16]. The categories of high, moderate, and low risk of bias will used based on the criteria developed by Grooten et al [16]: green (low risk of bias) for studies where all six QUIPs domains are assessed as low risk of bias (RoB) or not more than one domain is assessed as moderate risk of bias; red (high RoB) for studies where ≥ 1 is assessed as high RoB, or ≥ 3 domains are assessed as moderate RoB; yellow (moderate RoB) for all other articles in between. The same criteria will be used to assess the risk of bias rating for individual domains.

For the studies investigating the efficacy of fibrinogen administration, the Cochrane Risk of Bias tool will be used for Randomised control trials (RCTs) [17], and the ROBINS-I tool for non-randomised studies to determine the RoB [18]. The RoB of RCTs will be rated as high, low, or unclear. A green circle with a ‘+’ symbol within will be used for low RoB, a red circle with a ‘-’ symbol within for high RoB, and a yellow circle with a ‘?’ symbol within for unclear RoB. For the ROBINS-I tool the categories used will be low risk, moderate risk, serious risk, and critical risk of bias [18].

Publication bias will be assessed through funnel plots and Egger’s regression test, contingent on having an adequate number of studies. The GRADE framework will be used to assess the certainty of the evidence of the synthesis findings for each outcome measure in cases of fibrinogen administration and prognostication [19,20]. Two independent reviewers will conduct the GRADE assessments, with a third reviewer adjudicating any discrepancies.

If meta-analysis is not possible, the Synthesis without meta-analysis (SWiM) in systematic review reporting guidelines will be used to report narrative and qualitative data [21].

## Study Limitations

Due to the nature of this systematic review’s exclusion criteria, potentially valuable information in these excluded studies may be missed. Potentially valuable information from the data in studies that fail to report the events ratio or area under curve (AUC) whilst assessing the prognostic value of fibrinogen will also be missed as the data in these studies will not be included in our data analysis.

## Conclusions

Given the prevalence of TBI and the worldwide economic burden, optimising the care of TBI patients is pertinent. If fibrinogen has a prognostic value, it would help healthcare teams practise realistic medicine in accordingly adjusting the goals of care delivered to these patients based on their prognosis. If administering fibrinogen does indeed improve patient outcomes, then this would be an important treatment option for healthcare teams to consider incorporating for TBI patients, especially if fibrinogen administration is protective against haemorrhagic transformation of contusions for example.

Therefore, if we find that serum fibrinogen level has prognostic value and that administering fibrinogen improves outcome, this could be important in shared-decision making and in ensuring that patients have improved outcomes.

## Data Availability

All relevant data are within the manuscript and its Supporting Information files.

## Declarations

## Ethics and Study Dissemination

No ethical approval is required for this systematic review of already published data. The findings of this paper will be submitted for publishing in a peer-reviewed journal.

## Authors’ contributions

Joanne Igoli, Jeremiah Oluwatomi Itodo Daniel, and Temidayo Osunronbi contributed to the conception and design of the study. Joanne Igoli wrote the initial draft of the manuscript and is guarantor of the study. All authors contributed to subsequent versions of the manuscript. All authors read and approved the final manuscript.

## Acknowledgments

We would like to express our gratitude to the Surgery Interest Group of Africa for their support.

We would like to thank Idemudia Stephen Ogedegbe for his work on the protocol manuscript, after which his contribution ended.

